# A validated cloud-based genomic platform for co-ordinated, expedient global analysis of SARS-CoV-2 genomic epidemiology

**DOI:** 10.1101/2023.11.27.23298986

**Authors:** Daniel Gyamfi Amoako, Nguyen To Anh, Jasmine Bastable, Marc Brouard, Constanza Campano Romero, Andres Castillo Ramirez, Bede Constantinides, Derrick W. Crook, Phan Manh Cuong, Moussa Moise Diagne, Amadou Diallo, Nguyen Thanh Dung, Laura Dunn, Le Van Duyet, Josie Everatt, Katherine Fletcher, Philip W. Fowler, Mailie Gail, Jessica Gentry, Saheer Gharbia, Hospital for Tropical Diseases SARS-CoV-2 testing team, Nguyen Thi Thu Hong, Martin Hunt, Zamin Iqbal, Katie Jeffery, Dikeledi Kekana, Thomas Kesteman, Jeff Knaggs, Marcela Lopes Alves, Dinh Nguyen Huy Man, Amy J. Mathers, Nghiem My Ngoc, Sarah Oakley, Hardik Parikh, Tim E.A. Peto, T. Phuong Quan, Marcelo Rojas Herrera, Nicholas Sanderson, Vitali Sintchenko, Jeremy Swann, Junko Takata, Nguyen Thi Tam, Le Van Tan, Pham Ngoc Thach, Ndeye Marieme Top, Nguyen Thu Trang, Van Dinh Trang, Robert Turner, H. Rogier van Doorn, Anne von Gottberg, Jeremy Westhead, Nicole Wolter, Bernadette C. Young

**Author notes:** **Corresponding author:** Junko Takata, Nuffield Department of Medicine, University of Oxford, Oxford, OX3 9DU, 01865 220856. All authors listed alphabetically.

## Abstract

**Background:** Viral sequencing has made critical contributions to our understanding of and response to the COVID-19 pandemic, but sequencing capacity and bioinformatic expertise remain limited in many settings. This proof-of-principle study aimed to demonstrate the utility of a cloud-based sequencing analysis pipeline, the Tiled Amplicon Pipeline (TAP), for rapid and collaborative SARS-CoV-2 sequencing across seven globally distributed sites.

**Methods:** In this cross-sectional study from July to August 2022, seven international sites submitted all SARS-CoV-2 sequence data generated over a two-week period to our cloud platform. No patient identifying information was uploaded, and human reads were removed prior to upload to the cloud. Users could opt in to share sample information with collaborators via a tagging system. The pipeline performed sequence assembly, lineage identification and relatedness analysis.

**Results:** Seven sites contributed 5,432 sequences, of which 5,342 (98.3%) were from clinical samples and 90 (1.7%) were controls. 4,470/5,342 (83.7%) clinical samples had sufficient coverage for lineage assignment. Omicron lineages dominated, with BA.5, BA.4 and BA.2 comprising the vast majority, consistent with contemporary epidemiological observations at the time. Phylogenetic analysis demonstrated low diversity within lineages, and genotypically identical or highly similar sequences were recovered from globally disparate sites.

**Conclusions:** A cloud-based analysis platform like TAP addresses bioinformatic bottlenecks and facilitates international capacity building and collaboration in pathogen surveillance, enhancing global epidemic and pandemic preparedness.

**Importance:** The COVID-19 pandemic showed the potential of whole-genome sequencing in informing public health responses, but also highlighted the significant global disparity of sequencing capacity and bioinformatic expertise. In this study, we describe the development and validation of a cloud-based, automated sequencing analysis pipeline for SARS-CoV-2 – the Tiled Amplicon Pipeline (TAP). 5,432 sequences from clinical samples across six continents were collected over a two-week period and processed through TAP, demonstrating its power to co-ordinate secure data sharing and aggregated global analysis. TAP is designed to have the capability to assemble other viruses beyond SARS-CoV-2 that use tiled amplicon sequencing, highlighting its potential to be adapted quickly for use in future outbreaks. Tools such as TAP can simplify access to and interpretation of genomic data, which in turn contributes to a growing global landscape of genomic data sharing and surveillance that underpin pandemic preparedness.

## Introduction

Viral genome sequencing has proven pivotal to understanding the evolution of the SARS- CoV-2 virus during the COVID-19 pandemic and in shaping the public health response. International genomic surveillance and data sharing initiatives have together made it possible to track the emergence of variants globally [1], to demonstrate the impact of travel restrictions on viral dynamics across continents [2] or within countries [3, 4], and to identify transmission routes within hospitals [5]. Successive waves of infection driven by new variants showed that rapidly detecting new lineages is critical for understanding disease epidemiology and guiding subsequent public health responses [6], as well as informing the development of vaccines and therapeutics such as neutralising antibodies [7–9].

However, the pandemic also highlighted marked global variability in sequencing capacity and cost [10], with much of the SARS-CoV-2 sequencing undertaken at centralised reference laboratories [11]. Several important challenges remain in expanding genomic surveillance, including (1) access to sequencing technology, especially in lower- and middle-income countries, such as access to consumables, complex import requirements, or failure of supply chains; (2) availability of bioinformatic expertise; and (3) interpretability of results generated using a plethora of different wet lab and bioinformatics protocols [12–14]. There is a need for accessible genomic surveillance infrastructure that can be used by researchers, microbiologists, and clinicians from any location, to deliver an up-to-date, global perspective of genomic evolution of pathogens with epidemic or pandemic potential.

One solution to both the shortage of bioinformatic expertise and the lack of global interpretability of results is web-accessible analysis infrastructure [15]. The Global Pathogen Analysis Service (GPAS) was rapidly set up in 2021 in response to the COVID- 19 pandemic by the University of Oxford as a cloud-based, globally accessible web platform. GPAS provided fast and secure access to a comprehensive SARS-CoV-2 genomic analysis pipeline, delivering genome assembly, variant calling and lineage classification from raw sequence data. Users retained control of their data but could opt in to share their data to facilitate comparisons in a wider context, empowering laboratories to control their own analysis. GPAS was validated to UKAS ISO 15189:2012 standards in a UK tertiary hospital clinical microbiology laboratory, and was commissioned by the UK Health Security Agency (UKHSA) for over a year to support Pillar 1 national surveillance (testing in UKHSA laboratories and NHS hospitals, for healthcare workers and those with a clinical need) and various clinical trials. GPAS provided proof-of-principle that standard microbiology laboratories without bioinformatics expertise can generate timely outputs for local surveillance and automatically submit sequences to public repositories.

In 2024, GPAS was upgraded as the Tiled Amplicon Pipeline (TAP) with major enhancements to several software components. Since January 2025 TAP has been deployed as the SARS-CoV-2 pipeline on EIT Pathogena (version 1.2.0), a multi-pipeline genomic pathogen analysis platform that is free of charge for users in low- and middle-income countries and accessible in all settings with internet access [16]. This service demonstrates the potential of cloud-based platforms to overcome barriers to democratising effective genomic surveillance, making advanced genomic analysis available to resource-limited laboratories.

In this paper, we report the results of a collaborative, cross-sectional SARS-CoV-2 sequencing study across seven globally distributed sites, which demonstrate the utility of a cloud-based sequencing platform in providing a quality assured, rapid, and integrated global snapshot of viral diversity. We further describe the components of the pipeline and discuss its potential for rapid adaptation in response to future viral pandemics.

## Materials and Methods

### Pipeline development

TAP was deployed to a cloud platform and controlled via a Command Line Interface (CLI) tool. Full methodology is described in the Appendix (Supplementary Appendix A). At upload, a universally unique identifier (UUID) is generated and assigned to each sample. The mapping between the UUID and the user’s sample identifier is downloaded and only held by the user, and FASTQ header lines are also truncated, ensuring that no potentially personally identifiable information is transmitted to the cloud platform. Tight access control to data applies within TAP, and by default data is not shared with other users unless explicitly authorised by the data owners.

Following FASTQ file upload to the cloud, processing commences automatically with no further user input required. Any reads mapping to the human genome are first removed [17], reducing the risk of any human reads being retained. Genome assembly is performed by an amplicon-aware genome assembly tool Viridian v1.3.1 [18], which scaffolds per-amplicon *de novo* assemblies into a single whole genome consensus assembly in FASTA format. Viridian was configured to use the SARS-CoV-2 reference genome Wuhan-Hu-1 (MN908947.3) for assembly and variant calling [19], and a library of seven amplicon primer schemes (AmpliSeq v1; ARTIC versions 3, 4.1, 5.3.2 (400), 5.2.0 (1200); Midnight 1200; VarSkip v1a-2b). When a primer scheme is not specified by the user, Viridian can automatically infer the most likely primer scheme from within its library.

After assembly, amino acid mutations are identified using Nextclade [20], and Pango lineages are assigned with Pangolin version 4.3.1 [21]. Aligned sequences are compared using a novel algorithm, FindNeighbour5, which identifies single nucleotide variant (SNV) distances between sequences without conflicting variant calls (0 SNVs) as well as those differing by one, two and three SNVs [22, 23]. The main outputs from the pipeline are presented in an access-restricted user interface portal and downloadable as a summary file containing the lineage assignment, a list of related samples (according to permissions), and metrics of the genome assembly (e.g. coverage, mean depth, amplicon dropouts, etc). Detailed intermediate files such as VCF or FASTA files are also available for download.

### Pipeline validation

GPAS was validated for both Illumina and Oxford Nanopore Technologies (ONT) sequencing platforms in 2021, using various datasets including a ‘truth set’ of cultured SARS-CoV-2 samples sequenced with multiple platforms and library preparations, annotated with manually curated variant calls (described in further detail in [18] and [24]), in addition to community samples collected in Northumbria, UK. Overall, GPAS showed negligible adjusted false call rates (less than 1/100,000 nucleotides) with respect to the ‘truth set’, and had high concordance (less than 1.9/1,000,000 discordant events) with the ARTIC assembly pipeline in use at the time.

TAP was subsequently validated against the same ‘truth set’ of cultured SARS-CoV-2 samples, which achieved 100% concordance with expected Pango lineages using the same Pangolin version. Concordance of TAP and GPAS was further compared using data from the current study, which showed 99.4% concordance in lineage calls (using the same Pangolin and Viridian versions), and nucleotide call discordance of 0.59/1,000,000 sites. Further details of the validation and comparison processes are described in Supplementary Appendix B.

### Sample frame, sequencing, and upload

Between July and August 2022, seven sequencing centres participated in a two-week sequencing pilot across seven countries: Senegal; Chile; South Africa; New South Wales (NSW), Australia; Vietnam; United Kingdom (UK); and Virginia, United States (USA). Centres were either accredited clinical microbiology and/or public health laboratories. All clinical samples in which SARS-CoV-2 was detected by PCR in respiratory samples were eligible for inclusion and underwent in-house genomic sequencing at participating sites, with no more than one submission from an individual patient. Sequencing platform (either Illumina or ONT) and primer schemes were chosen by participating sites, who followed their established sequencing protocols.

Raw sequences were uploaded in FASTQ format to GPAS, the working name of the pipeline at the time, along with limited associated metadata (sample identifier, instrument platform, sample type [clinical or control], collection date, and country). Submitting sites verified the run quality reports of each sequencing batch including a review of positive and negative controls, and batches were passed or failed accordingly. All passed samples were tagged to release them to the shared data pool for subsequent aggregate analysis; explicit permission was given from all collaborators to configure data access controls such that all submitters could access and view each other’s sequences, metadata, and analytical outputs. For this study, automatic detection of primer scheme was enabled. All sequences were uploaded to the European Nucleotide Archive (project accession: PRJEB70597) at the time of the study.

### Data analysis

FASTQ files from the study were redownloaded from the ENA and run through EIT Pathogena (version 1.2.0 pre-release) on 6^th^ November 2024. A maximum likelihood phylogeny of aligned sequences was constructed for the three largest Pango lineages in the sample set (BA.5, BA.4 and BA.2) using IQTree version 2.3.6, assuming a general time reversible nucleotide substitution model with gamma rate heterogeneity [25]. All other analyses were conducted in RStudio version 2023.06.0+421.

## Results

### Primer scheme detection

5,432 sequences were shared across seven sites (Supplementary Table S1), with date of collection ranging from April to July 2022. Of submitted samples, 90 (1.7%) were controls and 5,342 (98.3%) were sequences from clinical samples. Primer schemes were successfully auto-detected by Viridian for six out of seven sites; for the remaining site (NSW, Australia), the primer scheme was not auto-detected as it used a modified Midnight-1200 scheme to account for amplicon dropouts, and was therefore explicitly specified.

### Aggregate analysis of global genetic epidemiology

Among the clinical samples included in the analysis, 4,782/5,342 (90.0%) were assembled with at least 70% genome coverage (Table 1). 4,470/5,342 (83.7%) clinical samples were assembled with sufficient coverage and post-assembly quality to be assigned a lineage (Table 2). 4,442/4,470 (99.4%) were Omicron variants, with BA.5, BA.4 and BA.2 being the most common. A small number of Delta variant sequences were identified (0.3%, 15/4,470), all of which were collected prior to July 2022.

**Table 1:** Samples submitted by study centre, with date of collection and genome coverage. Earliest and latest sample dates exclude controls. NSW = New South Wales. includes samples that could not be assembled

| **Centre** | **Submitted samples** | **Controls** | **<50% coverage\*** | **50-70% coverage** | **>70% coverage** | **Earliest sample collection date** | **Latest sample collection date** |
| --- | --- | --- | --- | --- | --- | --- | --- |
| **Senegal** | 197 | 0 | 9 | 10 | 178 | 13th July 2022 | 20th July 2022 |
| **Chile** | 1205 | 20 | 4 | 6 | 1175 | 16th June 2022 | 22nd July 2022 |
| **South Africa** | 202 | 0 | 7 | 6 | 189 | 17th May 2022 | 24th June 2022 |
| **NSW, Australia** | 1194 | 31 | 132 | 0 | 1031 | 21<sup>st</sup> June 2022 | 18<sup>th</sup> July 2022 |
| **Vietnam** | 316 | 3 | 14 | 7 | 292 | 1st April 2022 | 14th July 2022 |
| **United Kingdom** | 1818 | 36 | 280 | 85 | 1417 | 12th July 2022 | 26th July 2022 |
| **Virginia, USA** | 500 | 0 | 0 | 0 | 500 | 7th June 2022 | 28th June 2022 |
| **Total** | **5432** | **90** | **446** | **114** | **4782** |  |  |

**Table 2:** SARS-CoV-2 lineage of samples by study centre (where a lineage was assigned by Pangolin). NSW = New South Wales. Other includes: P.1, P.1.2, A, A.2.2, B.1, B.1.429, B.28, C.37, DE.1, XAM, XAN, XAS, XAZ, XBA, P.2

| Centre | Omicron |  |  |  |  |  |  | Delta | Alpha | Other* | Total |
| --- | --- | --- | --- | --- | --- | --- | --- | --- | --- | --- | --- |
|  | BA.1 | BA.2 | BA.4 | BA.5 and related |  |  |  | AY.5, AY.57, | B.1.1.7 |  |  |
|  |  |  |  | BA.5 | BE | BF | Other (BG, BK) | B.1.617.2 |  |  |  |
| Senegal | 0 | 17 | 24 | 28 | 2 | 12 | 0 | 0 | 0 | 0 | 83 |
| Chile | 0 | 136 | 570 | 302 | 26 | 63 | 1 | 0 | 0 | 5 | 1103 |
| South Africa | 0 | 3 | 82 | 77 | 8 | 3 | 0 | 0 | 0 | 3 | 176 |
| NSW, Australia | 0 | 165 | 124 | 629 | 58 | 52 | 2 | 0 | 0 | 1 | 1031 |
| Vietnam | 2 | 228 | 0 | 6 | 0 | 0 | 0 | 15 | 0 | 0 | 251 |
| United Kingdom | 2 | 57 | 165 | 864 | 137 | 111 | 0 | 0 | 1 | 2 | 1339 |
| Virginia, USA | 0 | 289 | 89 | 84 | 12 | 9 | 3 | 0 | 0 | 1 | 487 |
| Total | 4 | 895 | 1054 | 1990 | 243 | 250 | 6 | 15 | 1 | 12 | 4,470 |

### Global mixing across multiple Omicron lineages

For clinical samples collected between 1^st^ June 2022 and 31st July 2022, the proportion of samples assigned to the different Omicron sub-lineages varied substantially by study site (Figure 1). Each of BA.2, BA.4 or BA.5 dominated (constituted >50% of samples from) at least one site, while in Senegal no single lineage dominated, and BA.4 and BA.5 were equally common in South Africa (44.3% and 46.9% respectively).

**Figure 1:**
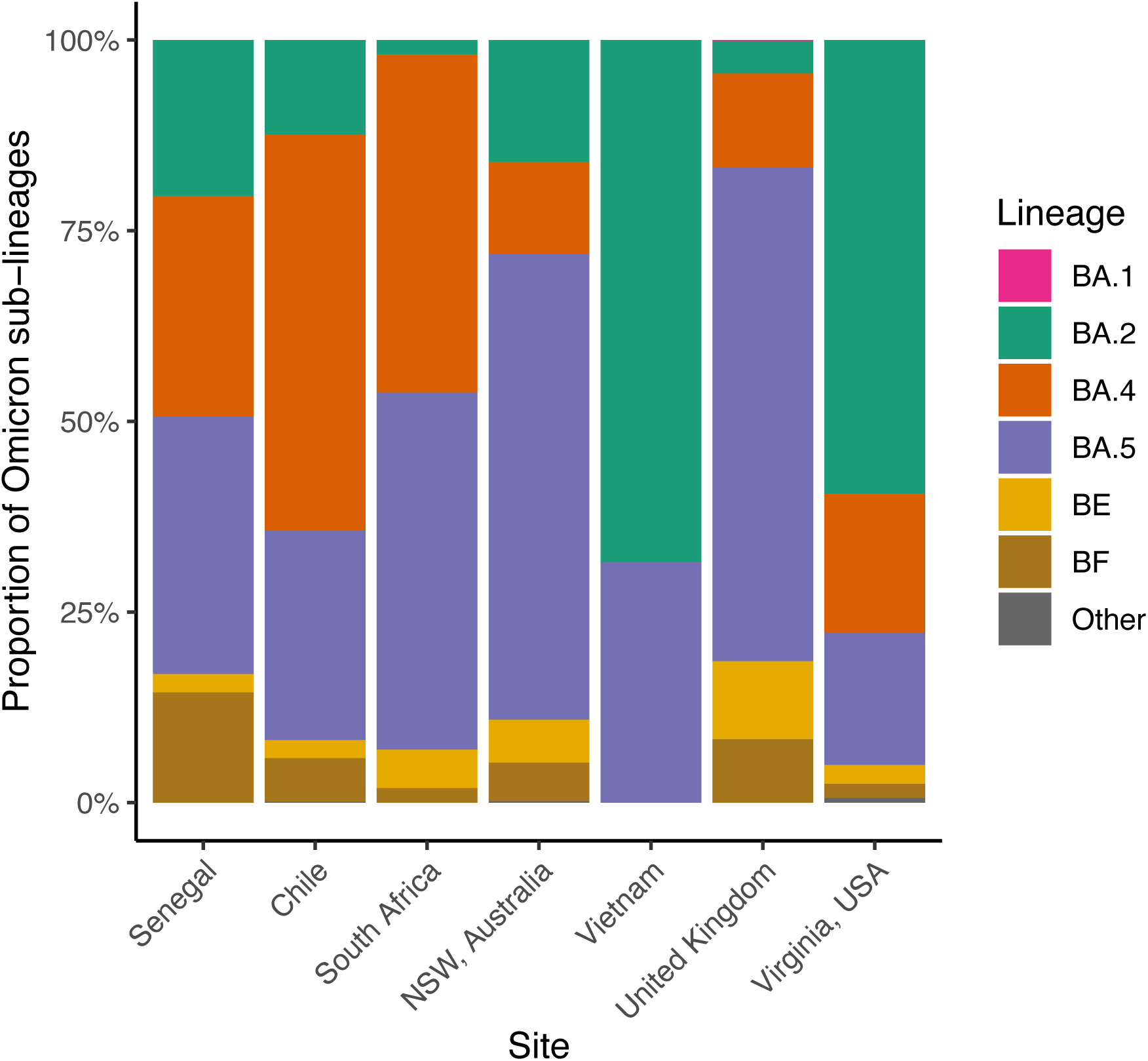
Omicron sub-lineages by study site. Proportion of samples assigned to Omicron sub-lineages within each study site, where collection date was in June or July 2022.

Maximum likelihood phylogenies for each of the three most common lineages revealed global mixing of Omicron lineages (Figure 2). There were no examples of sub-lineages being completely geographically restricted to one site; one sub-lineage of BA.4 was found predominantly in Chile, but an example of this sub-lineage was also identified in the United Kingdom.

**Figure 2:**
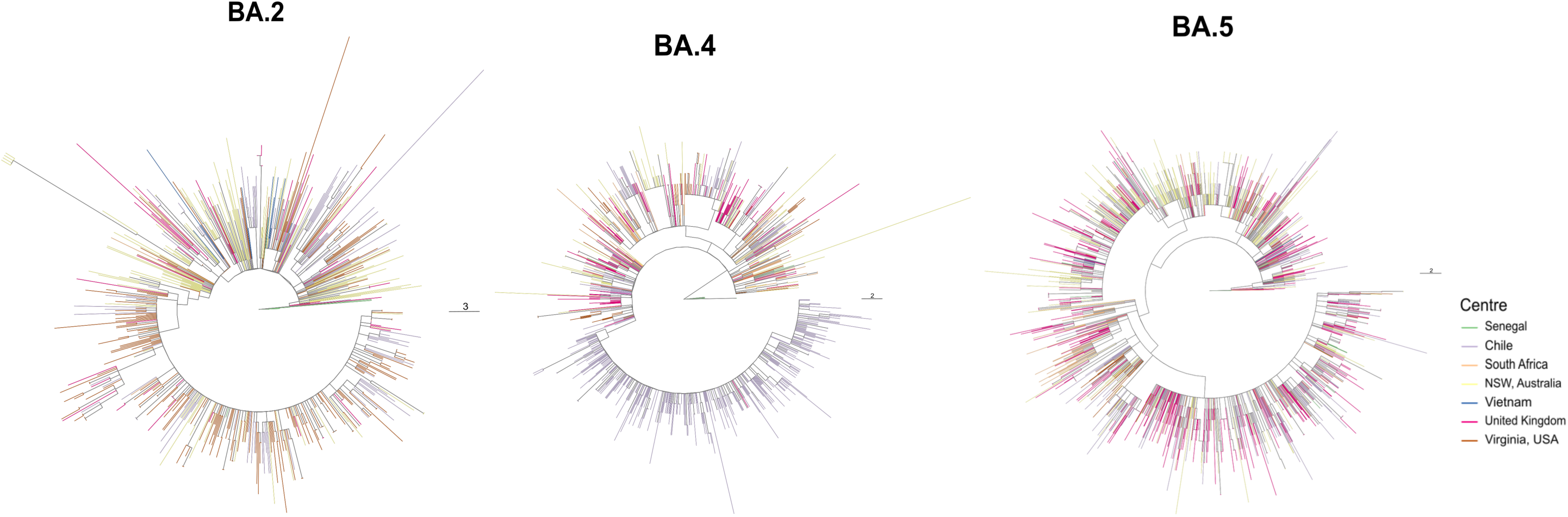
Maximum likelihood phylogeny of isolates from major SARS-CoV-2 lineages (BA.2, BA.4, BA.5), with branches coloured by sequencing site. Branch lengths are scaled to represent substitutions across the full SARS-CoV-2 genome with corresponding scale bars.

Similarly, a proportion of genotypically identical (SNV=0) isolate pairs were found in different sites for all 3 major lineages, particularly for the BA.2 lineage, where 21.3% of SNV=0 pairs were found in different sites (Figure 3, Supplementary Table 2). This proportion increased with greater SNV distances. BA.4 showed the least amount of global dispersal below SNV <3, corresponding to the majority of SNV <3 isolate pairs in this lineage being from Chile where there was evidence of a dominant sub-lineage as illustrated in Figure 2.

**Figure 3:**
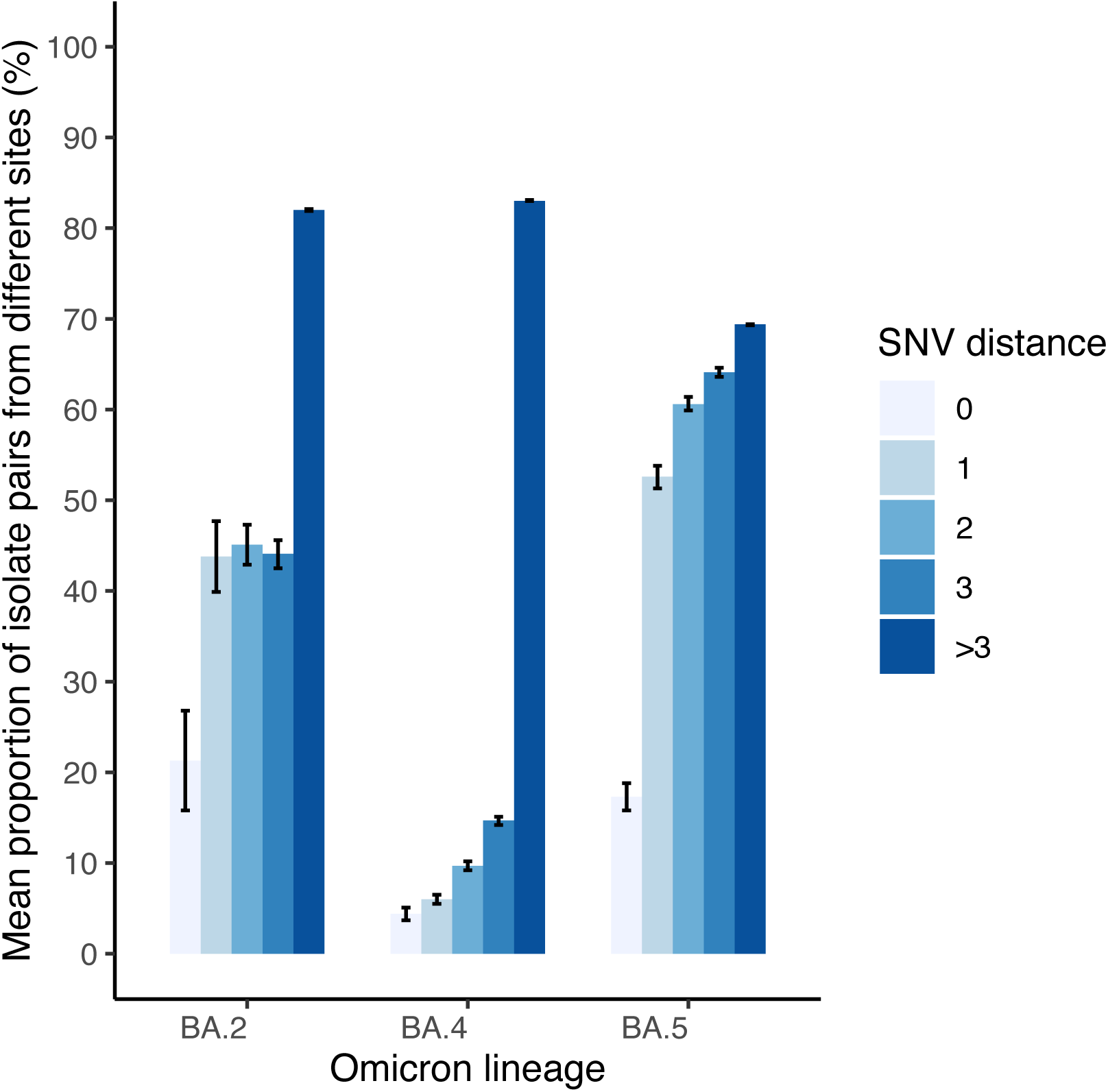
Proportion of unique isolate pairs that are from different sites at each SNV distance for each major Omicron lineage. Bars indicate 95% confidence intervals.

### Cloud processing time and performance

At the time of the study, cloud processing performance on GPAS was measured by time elapsed between upload and completion of analysis. Median cloud processing time per sample was 30.6 minutes (IQR: 13.1 – 69.9); variability was observed, with several batches from Australia, UK and USA taking longer than 1000 minutes (Supplementary Figure 1). These were caused by either a data centre outage during an extreme heat event in the UK, or as a result of a software update prematurely applied to the upload portal, which was promptly resolved by improvements in the client side CLI software. Other data upload issues were identified and addressed case-by-case, including metadata file formatting errors, the availability of upload clients compatible with all required operating systems, and user interface display errors preventing batch release.

In TAP, median cloud processing time per sample for a typical batch of 100 was 14.5 minutes (IQR: 13.8 – 15.0) for Illumina and 15.6 minutes (IQR: 15.2 – 16.7) for ONT; due to parallelisation, processing a single batch of 100 samples was achievable in 19.7 minutes (Illumina) and 20.4 minutes (ONT) (Supplementary Figure 2). Processing all 5,432 samples took 766 minutes (12.7 hours). CPU time per sample is approximately 3 minutes (noting that this occurs in parallel, i.e. 2 CPUs working for 4 minutes would be reported as 8 minutes CPU time by the workflow manager, despite only taking 4 minutes); majority of analysis time is associated with other pipeline activities including network file transfers or reading data from disk into memory. Peak RAM usage for typical use of the pipeline was approximately 5GB.

## Discussion

This pilot study demonstrates the potential of globally synchronous data processing and analysis in informing public health, through a unified protocol of genome assembly, variant calling and relatedness analysis. The aggregated analysis from this study conforms to contemporary observations of SARS-CoV-2 genetic epidemiology at the time [26–28], and demonstrates cosmopolitan global mixing of Omicron lineages between study sites. Even a BA.4 sub-lineage found almost exclusively in Chile was not completely restricted to that site and could be identified in a geographically distant site (UK). Similarly, up to 21% of genotypically identical (SNV=0) sequence pairs were identified in different countries over the same two-month period. Such mixing of lineages likely reflects the relaxation of travel restrictions in the participating sites at the time of the study, and captures the rapid global dispersal of Omicron with successive selective sweeps of new lineages over short time scales [29]. These observations highlight the potential of a cloud-based sequencing pipeline in facilitating data sharing and generating co-ordinated insights that could inform real-time decision-making.

While automatic primer scheme detection was used for this study, our results show the limitation in this approach, where a modified scheme was not detected to be amongst the current Viridian primer scheme library. Although this illustrates the sensitivity of Viridian’s auto-detection, it is inevitable that with viral evolution, primer schemes will continue to be modified or newly developed to maintain sequencing coverage of an evolving target genome. Viridian has the capability to accommodate custom primer schemes when provided with a suitable scheme definition in browser extensible data (BED) format. This capability will therefore form the basis of future pipeline iterations and will allow the pipeline to pivot in future to assemble other viruses beyond SARS-CoV-2 that use tiled amplicon sequencing. In turn, this ability to rapidly adapt existing infrastructure for emerging threats, rather than build bespoke solutions from scratch each time, supports a proactive and ‘Always On’ approach to pandemic preparedness [30].

Our study is limited by the opportunistic sampling frame used and is likely to be geographically incomplete, with a preponderance for countries that have historically contributed more to global sequencing efforts. Similarly, our study has the limitation of lacking longitudinal data on how these findings changed over time. Despite these shortcomings, we have demonstrated how even a simply structured sampling across time and space is still well placed to rapidly identify replacement by new lineages. Such limitations in sampling frame diversity and global disparity in sequencing capacity were reflected during the pandemic itself: sequencing data produced in high-income countries was up to ten-fold higher than that achieved in low- and middle-income countries during the first two years [11], and data was not collected evenly over the course of pandemic. Well-designed, suitably powered, and representative sampling is a key part of national and international pandemic preparedness planning, as exemplified by a cohort design in the UK [27].

The world is now better prepared to urgently initiate genomic pathogen surveillance. The available sequencing platform capacity is much improved, and the achievements of global data aggregation (as illustrated by INSDC, GISAID and Pathoplexus databases) have alleviated a major obstacle to effective genomic surveillance and data sharing. In turn, the integration of genomic data with disease manifestation and severity data would enable the ready investigation of associations between genomic variation and clinical outcome as suggested by others [31], and facilitate predictive modelling for anticipating the course of future epidemics [32].

A remaining obstacle, especially in low-resource settings, is establishing local turnkey bioinformatics to ensure standardised, quality-assured outputs that do not depend on in-house bioinformatics expertise. In TAP, we have developed an exemplar of such a cloud-based service, with simple ingestion of local, raw sequence data in FASTQ format, and flexible privacy protections for data sharing to facilitate local or international comparative analyses. Outputs from TAP such as the consensus FASTA sequence can in turn be submitted to international repositories such as GISAID which do not take raw FASTQ files directly. While factors such as internet bandwidth may limit real-world performance, TAP is capable of rapid parallel data processing, with a run time of 15 minutes per sample and 20 minutes in total for a batch of 100 typical SARS-CoV-2 genomes. The EIT Pathogena platform ensures sustainability of technical and infrastructural support to the pipeline and is currently freely available to low- and middle-income countries, supporting long-term use across research, clinical, or surveillance settings. Such a service, in turn, contributes to the growing global landscape of genomic data sharing that will underpin future pandemic preparedness.

## Conclusions

The COVID-19 pandemic has demonstrated the multiple ways in which viral sequencing can inform pandemic responses, but also the inequitable distribution of resources and capabilities. A service such as TAP contributes to the democratisation of genomics, enabling researchers and laboratories, regardless of their location or bioinformatics expertise, to participate actively in global surveillance efforts. By providing accessible and user-friendly tools for sequence assembly, lineage assignment, and data sharing, it promotes inclusive collaboration, harmonises data, and ultimately enhances pandemic preparedness.

## Author contributions

Study conceptualisation: BCY, DWC

Study design: BCY, DWC

Software building and computational infrastructure: MB, BC, MH, ZI, PWF, JK, MLA, NS, JS, RT, JW

Data collection: DA, NTA, JB, CCR, ACR, PMC, MMD, AD, NTD, LD, LVD, JE, KF, MG, JG, SG,

Hospital for Tropical Diseases SARS-CoV-2 testing team, NTTH, KJ, DK, TK, DNHM, AJM, NMN, SO, HP, MRH, VS, NTTa, LVT, PNT, NMT, NTTr, VDT, HRVD, AvG, NW

Data analysis: JT, BCY, BC, TEAP, TPQ Manuscript writing: JT, BCY, BC Manuscript editing and review: all authors

## Ethical statement

SARS-CoV-2 sequencing was performed for public health surveillance and ethical approval for secondary analysis was not required. This determination was reviewed by the University of Oxford Joint Research Office. No patient identifying information was shared as part of this study. Sample identifiers remain with the submitter, and are never kept on GPAS/TAP, or shared with other users. GPAS/TAP generates identifiers for each sequence and writes a file linking anonymised ID to the submitted identifiers. Only the user submitting sequence data has access to these records. The GPAS/TAP upload client selects only SARS-CoV-2 sequence reads and discards all human sequences.

## Data availability

Submitted sequencing reads (free from human reads) for all included samples are available from European Nucleotide Archive study accession PRJEB70597.

## Acknowledgments

Microbial Genomics Reference Laboratory, New South Wales Health Pathology, Sydney, Australia

- El Instituto de Salud Pública de Chile, Chile
- Centre for Respiratory Diseases and Meningitis (CRDM), National Institute for Communicable Diseases (NICD), a division of the National Health Laboratory Service, South Africa
- The Institut Pasteur de Dakar, Senegal
- Oxford University Hospitals NHS Trust, Oxford, United Kingdom
- University of Virginia, Charlottesville, United States of America
- Oxford University Clinical Research Unit, Vietnam
- Hospital for Tropical Diseases SARS-CoV-2 testing team, HTD Vietnam: Le Manh Hung, Nguyen Le Nhu Tung, Nguyen Thanh Phong, Vo Minh Quang, Pham Thi Ngoc Thoa, Nguyen Thanh Truong, Tran Nguyen Phuong Thao, Dao Phuong Linh, Ngo Tan Tai, Ho The Bao, Vo Trong Vuong, Huynh Thi Kim Nhung
- Oracle Global Health Business Unit

## References

1. Li J, Lai S, Gao GF, Shi W. The emergence, genomic diversity and global spread of SARS-CoV-2. Nature. 2021;600(7889):408-18.

2. Wilkinson E, Giovanetti M, Tegally H, San JE, Lessells R, Cuadros D, et al. A year of genomic surveillance reveals how the SARS-CoV-2 pandemic unfolded in Africa. Science. 2021;374(6566):423-31.

3. McCrone JT, Hill V, Bajaj S, Pena RE, Lambert BC, Inward R, et al. Context-specific emergence and growth of the SARS-CoV-2 Delta variant. Nature. 2022;610(7930):154- 60.

4. Raghwani J, du Plessis L, McCrone JT, Hill SC, Parag KV, Theze J, et al. Genomic Epidemiology of Early SARS-CoV-2 Transmission Dynamics, Gujarat, India. Emerg Infect Dis. 2022;28(4):751-8.

5. Snell LB, Fisher CL, Taj U, Stirrup O, Merrick B, Alcolea-Medina A, et al. Combined epidemiological and genomic analysis of nosocomial SARS-CoV-2 infection early in the pandemic and the role of unidentified cases in transmission. Clin Microbiol Infect. 2022;28(1):93–100.

6. Updated working definitions and primary actions for SARS-Cov-2 variants. WHO Technical Advisory Group on Virus Evolution; 2023.

7. Tao K, Tzou PL, Nouhin J, Gupta RK, de Oliveira T, Kosakovsky Pond SL, et al. The biological and clinical significance of emerging SARS-CoV-2 variants. Nat Rev Genet. 2021;22(12):757–73.

8. Tuekprakhon A, Nutalai R, Dijokaite-Guraliuc A, Zhou D, Ginn HM, Selvaraj M, et al. Antibody escape of SARS-CoV-2 Omicron BA.4 and BA.5 from vaccine and BA.1 serum. Cell. 2022;185(14):2422-33 e13.

9. Aggarwal A, Akerman A, Milogiannakis V, Silva MR, Walker G, Stella AO, et al. SARS-CoV-2 Omicron BA.5: Evolving tropism and evasion of potent humoral responses and resistance to clinical immunotherapeutics relative to viral variants of concern. EBioMedicine. 2022;84:104270.

10. Merhi G, Koweyes J, Salloum T, Khoury CA, Haidar S, Tokajian S. SARS-CoV-2 genomic epidemiology: data and sequencing infrastructure. Future Microbiol. 2022;17:1001–7.

11. Brito AF, Semenova E, Dudas G, Hassler GW, Kalinich CC, Kraemer MUG, et al. Global disparities in SARS-CoV-2 genomic surveillance. Nat Commun. 2022;13(1):7003.

12. Carter LL, Yu MA, Sacks JA, Barnadas C, Pereyaslov D, Cognat S, et al. Global genomic surveillance strategy for pathogens with pandemic and epidemic potential 2022-2032. Bull World Health Organ. 2022;100(4):239-A.

13. Inzaule SC, Tessema SK, Kebede Y, Ogwell Ouma AE, Nkengasong JN. Genomic- informed pathogen surveillance in Africa: opportunities and challenges. Lancet Infect Dis. 2021;21(9):e281–e9.

14. Chen Z, Azman AS, Chen X, Zou J, Tian Y, Sun R, et al. Global landscape of SARS- CoV-2 genomic surveillance and data sharing. Nat Genet. 2022;54(4):499–507.

15. Ohlsen EC, Hawksworth AW, Wong K, Guagliardo SAJ, Fuller JA, Sloan ML, et al. Determining Gaps in Publicly Shared SARS-CoV-2 Genomic Surveillance Data by Analysis of Global Submissions. Emerg Infect Dis. 2022;28(13):S85–S92.

16. EIT Pathogena [Available from: https://eit-pathogena.com.

17. Constantinides B, Hunt M, Crook DW. Hostile: accurate decontamination of microbial host sequences. Bioinformatics. 2023;39(12).

18. Hunt M, Hinrichs AS, Anderson D, Karim L, Dearlove BL, Knaggs J, et al. Addressing pandemic-wide systematic errors in the SARS-CoV-2 phylogeny. bioRxiv. 2024:2024.04.29.591666.

19. Wu F, Zhao S, Yu B, Chen YM, Wang W, Song ZG, et al. A new coronavirus associated with human respiratory disease in China. Nature. 2020;579(7798):265-9.

20. Aksamentov I, Roemer C, Hodcroft EB, Neher RB. Nextclade: clade assignment, mutation calling and quality control for viral genomes. Journal of Open Source Software. 2021;6(67):3773.

21. O’Toole A, Scher E, Underwood A, Jackson B, Hill V, McCrone JT, et al. Assignment of epidemiological lineages in an emerging pandemic using the pangolin tool. Virus Evol. 2021;7(2):veab064.

22. Mazariegos-Canellas O, Do T, Peto T, Eyre DW, Underwood A, Crook D, et al. BugMat and FindNeighbour: command line and server applications for investigating bacterial relatedness. BMC Bioinformatics. 2017;18(1):477.

23. FindNeighbour5 [Available from: https://github.com/oxfordmmm/FN5.

24. Constantinides B, Webster H, Rodger G, Hunt M, Supasa P, Dejnirattisai W, et al. A diverse reference set of cultured SARS-CoV-2 genomes sequenced using various amplification methods and instrument platforms 2024 [Available from: https://www.ebi.ac.uk/biostudies/studies/S-BSST1334.

25. Minh BQ, Schmidt HA, Chernomor O, Schrempf D, Woodhams MD, von Haeseler A, et al. IQ-TREE 2: New Models and Efficient Methods for Phylogenetic Inference in the Genomic Era. Mol Biol Evol. 2020;37(5):1530–4.

26. Tosta S, Moreno K, Schuab G, Fonseca V, Segovia FMC, Kashima S, et al. Global SARS-CoV-2 genomic surveillance: What we have learned (so far). Infect Genet Evol. 2023;108:105405.

27. Foulkes S, Monk EJM, Sparkes D, Hettiarachchi N, Milligan ID, Munro K, et al. Early Warning Surveillance for SARS-CoV-2 Omicron Variants, United Kingdom, November 2021-September 2022. Emerg Infect Dis. 2023;29(1):184-8.

28. Xu Y, Liu T, Li Y, Wei X, Wang Z, Fang M, et al. Transmission of SARS-CoV-2 Omicron Variant under a Dynamic Clearance Strategy in Shandong, China. Microbiol Spectr. 2023;11(2):e0463222.

29. Markov PV, Ghafari M, Beer M, Lythgoe K, Simmonds P, Stilianakis NI, et al. The evolution of SARS-CoV-2. Nat Rev Microbiol. 2023;21(6):361–79.

30. van der Westhuizen HM, Soundararajan S, Berry T, Agus D, Carmona S, Ma P, et al. A consensus statement on dual purpose pathogen surveillance systems: The always on approach. PLOS Glob Public Health. 2024;4(11):e0003762.

31. Oude Munnink BB, Worp N, Nieuwenhuijse DF, Sikkema RS, Haagmans B, Fouchier RAM, et al. The next phase of SARS-CoV-2 surveillance: real-time molecular epidemiology. Nat Med. 2021;27(9):1518–24.

32. Volz E, Mishra S, Chand M, Barrett JC, Johnson R, Geidelberg L, et al. Assessing transmissibility of SARS-CoV-2 lineage B.1.1.7 in England. Nature. 2021;593(7858):266- 9.

