## Supplementary Appendix for "A validated cloud-based genomic platform for co-ordinated, expedient global analysis of SARS-CoV-2 genomic epidemiology"

### 1 Supplementary Appendix A: Pipeline overview

#### 1.1 Overview

The Tiled Amplicon Pipeline was deployed on Oracle Cloud Infrastructure and configured using Terraform. Nextflow is used to standardise the pipeline steps and manage sequencing of tasks. A summary of a similarly engineered pipeline for mycobacteria was described previously [1]. The pipeline was optimised for SARS-CoV-2 and it is described in those terms below.

After sample upload, human read removal is performed in the cloud and the original upload sample is deleted. The amplicon scheme used for tiling PCR amplification is either selected by the user or automatically detected, following which reads are assembled against the reference SARS-CoV-2 genome, mutations called, and Pango lineage assigned. Relatedness in terms of SNV distance between the sample and others in the database is called. Outputs from the pipeline are drawn together in a summary step, and files generated at each stage of the pipeline are downloadable by the user. The pipeline is available both via a Command Line Interface (CLI) and a web browser user interface.

Input

Human Read

Removal

Species Check

Genome assembly

Lineage assignment

Mutation calling

Summary

Relatedness

Database

**Pipeline Overview**. Following human read removal and genome assembly, the pipeline goes through three parallel steps of lineage assignment, mutation calling and relatedness. Outputs from the lineage assignment and mutation calling are passed to a summary file, while outputs from the relatedness analysis is displayed separately to the user. A check to ensure the correct species runs in parallel.

#### 1.2 Human Read Removal: Hostile

Human read removal is the process of removing human reads from FASTQ files. Here we use Hostile [2]. The process carried out is:

1. Reads are aligned to a custom human reference genome using Minimap 2 [3] for Oxford Nanopore Technologies (ONT) reads or Bowtie 2 [4]) for Illumina reads.
2. Distinct reads are counted using SAMTools [5] so that the number of reads removed can be subsequently calculated.
3. Reads that align are discarded (along with their mate reads for paired data) using SAMTools.
4. Remaining reads are counted using SAMTools.
5. Optionally, read names are replaced with incrementing integers (Awk), and (vi).
6. Outputs are written as gzip-compressed FASTQ files using SAMTools.

#### 1.3 Species Check: Gatekeeper

Our implementation of read quality checks is called Gatekeeper. Currently, Kraken 2 [6] is used as a check to ensure that all species detected in the sample are consistent with the intended species. This allows diagnosis of a potential reason in cases of failure to assemble a genome.

#### 1.4 Genome assembly: Viridian

Genome assembly is performed by Viridian v1.3.1, a genome assembly tool designed for tiled amplicon sequencing data, described in detail elsewhere [7]. In brief:

1. The amplicon scheme is identified. Viridian can automatically detect the best matching scheme from the available list of built-in schemes, but this can also be specified by the user from the available list. For SARS-CoV-2, the pipeline currently supports AmpliSeq v1; ARTIC versions 3, 4.1, 5.3.2 400, 5.2.0 1200; Midnight 1200; and VarSkip v1a-2b. Custom amplicon schemes can also be added to Virdian if a BED file is supplied, but this is not currently a feature of our pipeline.
2. At each amplicon level, reads are sampled to a target depth of 100X (or all reads if mean depth is less than 100X), and a consensus sequence per amplicon is generated.
3. The per-amplicon consensus sequences are overlapped into contigs, and mapped to a reference genome to order and orient them to create an overall consensus sequence. A whole genome assembly in FASTA format is produced by Viridian, and used for lineage calling and mutation analysis. From this assembly, a fixed length FASTA is derived for use in subsequent relatedness analysis.

Quality control is performed at each level.

- An amplicon is failed if its consensus sequence is shorter than 30bp or has more than 50% Ns. The pipeline is stopped if less than 50% of the genome has at least 20X read depth, and/or if greater than 50% of amplicons are dropped.
- Automatic amplicon detection fails if the best matching amplicon scheme has a score less than two-fold as high as the second-best amplicon scheme (max scheme ratio of 0.5).
- Low quality positions, where the majority of reads disagree with the consensus or fewer than 20 reads agree with the consensus, are masked. Positions with a mix of nucleotides supported by the reads are replaced with ambiguous IUPAC codes – these are referred to as ‘heterozygous’ calls.
- Following genome assembly, assemblies with more than 5,000 N calls and more than 3 heterozygous base calls are filtered out from subsequent steps, in accordance with the quality control measures used in the Viridian global tree [7].

#### 1.5 Lineage assignment: Pangolin

Pangolin v4.3.1 [8] is used for assigning lineage names to SARS-CoV-2 genomes according to the Pango nomenclature system [9]. Input sequences are aligned against an early SARS-CoV-2 reference sequence, and Scorpio is used to identify lineages corresponding to defined variants of concern. This is integrated with the outputs from a parallel inference pipeline using UShER, and merged to call the final lineage.

#### 1.6 Mutation calling: Nextclade and BCFtools

The assembled genome is compared against the reference by Nextclade, which calls mutations at a nucleotide and amino acid level, and by BCFTools, which is used for consequence calling (e.g. synonymous or nonsynonymous mutation). This information is included in the final summary report, but was not directly used in the analysis described in this paper.

#### 1.7 Relatedness: FindNeighbour5

Relatedness is measured as calculation of SNV (single nucleotide variant) distance from a fixed length FASTA file with our in-house algorithm FindNeighbour5 [10]. For each new sample processed:

1. The FASTA file of the reference genome is read.
2. A mask is used to ensure repetitive or highly variable regions are ignored. The choice of mask positions for SARS-CoV-2 is defined in [11], and includes problematic sites that are, for example, highly ambiguous or homoplasic.
3. The FASTA file of the sample is parsed and compared with the reference genome. If the sample is not identical to the reference at an unmasked base, the position is stored (along with a sample identifier). Non-ACGT nucleotides (i.e. IUPAC ambiguity codes) are treated equivalent to N.
4. These positions (arranged by nucleotide) are saved to disk.
5. The saves for each pre-existing sample are then loaded from disk, comparing SNV distance for each against the new sample. If the distance is above a defined cutoff (set as 3 for SARS-CoV-2), the distance calculation stops early (for efficiency). Where one sample is N (or a non-ACGT ambiguous nucleotide), a distance is not calculated.
6. The new sample is then added to the list of samples for future comparisons. This is performed in an asynchronous, distributed and thread-safe manner to ensure that distances are added appropriately.

Note that FN5 treats non-ACGT nucleotides equivalent to N, which may not be the case in other lineage assignment or phylogenetic algorithms. Therefore, a result of SNV=0 from FN5 does not necessarily indicate exactly identical Pango lineages, if, for example, a lineage-defining SNV is at an ambiguous site.

#### 1.8 Summary

The summary task brings together the outputs from other tasks in the pipeline into a single summary json file. Some outputs are subsequently pulled together into a database.

### 2 Supplementary Appendix B: Pipeline validation

#### 2.1 GPAS validation

The legacy platform (GPAS) was originally created during the COVID-19 pandemic in 2021 and was validated at that time to UKAS standards to support 'Pillar 1' national surveillance and clinical trials.

**2.1.1	Nucleotide false calls compared to a manually curated truth set**

Performance of GPAS was compared to ncov2019-artic-nf [12], a mapping-based assembly pipeline that was in use in 2021, using a reference set of 46 manually curated samples (‘truth set’). 16 of these samples were sequenced from viral culture at the University of Oxford, using ARTIC v3 primers with both Illumina and ONT (described in further detail under “Empirical truth set” in [7] and available on ENA study accession PRJEB51850). The other 30 samples were taken from publicly available clinical samples sequenced using ARTIC v3 with both Illumina and ONT at Johns Hopkins University, USA (described in further detail in [13] and available on ENA study accession PRJNA650037). VCF files for all 46 samples were manually inspected and variant calls curated using the methods described in [7].

Each sequence is 29,903 nucleotides long, providing 46*29,903 = 1,375,538 ‘truth set’ nucleotide sites; of this, 1,374,667 were the same as the wild-type Wuhan strain.

| **Truth assignment compared to wild type** | **Number of positions** |
| --- | --- |
| Het/unsure | 23 |
| Deletions | 201 |
| Substitutions | 647 |
| Wild type nucleotides | 1,374,667 |
| TOTAL | 1,375,538 |

Nucleotide calls made by GPAS and by ncov2019-artic-nf were compared to these ‘truth set’ nucleotide calls (excluding deletion and het/unsure sites in the ‘truth set’ vis-à-vis the wild-type Wuhan strain). Comparisons were stratified by instrument platform (Illumina and ONT); false calls indicate where a pipeline called a different nucleotide to the ‘truth set’ assignment.

False calls were negligible for substituted nucleotides. For wild type nucleotides, 30/32 of all the identified false calls were called the same by all 4 (except 1 call which was not called in Artic-ONT), suggesting that the truth set assignment was wrong. False call rates are therefore presented as both crude total false calls, as well as total ‘adjusted’ false calls where these 30 calls (or 29 calls in the case of Artic-ONT) are removed. Adjusted false call rates were below 1/100,000 in all 4 assembler-instrument combinations, with no significant difference between them (p=0.60).

|  | **Substitutions in truth set (n=647)** | | | **Wild type nucleotides in truth set (n=1,374,667)** | | | **Total false calls** | **Total adjusted false calls** | **Missing** |
| --- | --- | --- | --- | --- | --- | --- | --- | --- | --- |
|  | **True calls** | **False calls** | **Missing** | **True calls** | **False calls** | **Missing** | **n (rate per 100,000)** | **n (rate per 100,000)** | **n (%)** |
| **GPAS-Illumina** | 647 | 0 | 0 | 1373878 | 32 | 757 | 32 (2.33) | 2 (0.15) | 0.06 |
| **GPAS-ONT** | 639 | 8 | 0 | 1354676 | 30 | 19961 | 38 (2.80) | 8 (0.59) | 1.45 |
| **Artic-Illumina** | 646 | 1 | 0 | 1370364 | 30 | 4273 | 31 (2.26) | 1 (0.07) | 0.31 |
| **Artic-ONT** | 641 | 6 | 0 | 1367241 | 29 | 7397 | 35 (2.56) | 6 (0.44) | 0.54 |

**2.1.2	Nucleotide call concordance with ARTIC assembler**

Concordance of nucleotide calls made by GPAS and by the ARTIC assembler were compared using 147 samples.

135 samples were ‘community’ clinical samples collected in Northumbria, UK in 2021. In brief, RNA was extracted, amplified with ARTIC v4.1 primers, and sequenced with Illumina in Northumbria; an aliquot of the extracted RNA was amplified with Artic v4.1 primers and sequenced with ONT in Oxford. Of 178 initial samples, 43 had coverage too poor for GPAS to yield FASTA files, leaving 135 samples that had FASTA files in both assemblers. Consensus coverage varied considerably between samples; average coverage was similar in both assemblers (97.1% in GPAS, 98.4% in ARTIC).

12 samples were sequenced from viral culture at the University of Oxford (cultured and manually curated the same as the 16 samples in section 2.1.1; available on the ENA study accession PRJEB50520 and described in [14]). These samples were selected from a range of lineages (B.28, B, B.1, B.1.1.7, B.1.351, P.1, P.2, B.1.525, B.1.1.7_E484K, B.1.214.2, B.1.617.2, BA.1) and were sequenced four-ways (Illumina ARTIC V4.1, ONT ARTIC v4.1, ONT Midnight-1200 v2, ONT SISPA). For this analysis, the Illumina ARTIC v4.1 run and ONT Midnight-1200 v2 run were used.

Across all 4 datasets (135 community samples and 12 cultured ‘truth’ samples, each sequenced with Illumina and ONT), there were only 16 discordant events between GPAS and the ARTIC assembler.

| **Data source** | **Platform** | **Concordant sites between ARTIC and GPAS*** | | | | **Discordant sites between ARTIC and GPAS**  **sites (events)** | **Total sites ***** |
| --- | --- | --- | --- | --- | --- | --- | --- |
|  |  | **Wild type** | **Substitutions** | **Deletions ****  **sites (events)** | **Insertions ****  **sites (events)** |  |  |
| Cultured (n=12) | Illumina | 356,324 | 290 | 250 (29) | 24 (3) | 0 | 356,617 |
| Cultured (n=12) | ONT | 355,344 | 293 | 194 (30) | 24 (3) | 0 | 355,640 |
| Community (n=135) | Illumina | 3,919,395 | 5,304 | 1797 (408) | 3 (1) | 10 (10) | 3,924,710 |
| Community (n=135) | ONT | 3,857,736 | 5,230 | 1939 (396) | 2 (1) | 34 (6) | 3,863,001 |
| **Total (n=294)** |  | **8,488,799** | **11,117** | **4180 (863)** | **53 (8)** | **44 (16)** | **8,499,968** |

* Concordant sites are further divided into wild-type, substitutions, deletions or insertions in reference to the Wuhan ‘wild type’ strain. Nucleotides that were non-ACGT were not considered, hence, total number of nucleotides per row vary.

** Deletions and insertions are represented as both the number of sites involved as well as the number of events (e.g. 1 deletion event could involve 3 nucleotide sites, etc).

*** Total sites do not include deleted sites.

All 16 discordant events were found in community samples, with breakdown in relation to the Wuhan wild type reference strain shown below, showing discordant events are distributed across different discordance types. Overall, this corresponds to 16 discordant events out of 8,499,968 nucleotides, i.e. a discordant rate of 1.9/1,000,000 nucleotides.

| **Nucleotide call compared to wild type** | | **Instrument** | | **Total** |
| --- | --- | --- | --- | --- |
| **Artic** | **GPAS** | **Illumina** | **ONT** |  |
| Wild type | Substitution | 0 | 0 | 0 |
| Substitution | Wild type | 9 | 0 | 9 |
| Wild type | Insertion | 1 | 2 | 3 |
| Insertion | Wild type | 0 | 1 | 1 |
| Wild type | Deletion | 0 | 3 | 3 |
| Deletion | Wild type | 0 | 0 | 0 |
| **Total** |  | **10** | **6** | **16** |

**2.1.3	Lineage call concordance with ARTIC assembler**

Finally, concordance of lineage calls made by GPAS and by ARTIC were compared for the 135 ‘community’ samples (each with Illumina and ONT). Of 270 sequences, Nextclade outputs were identical in all samples. Pangolin lineages differed in 5 samples (sample 1-4, and sample 7 below) and no lineage was detected in GPAS in 4 samples (samples 5-8). Samples with lineage discrepancies (shown in red below) had high numbers of amplicon dropouts (mean dropout of 27/99 amplicons).

| **Sample** | **Instrument** | **NextClade** | | **Pangolin** | | **Scorpio** | | **Amplicons*** |
| --- | --- | --- | --- | --- | --- | --- | --- | --- |
|  |  | **ARTIC** | **GPAS** | **ARTIC** | **GPAS** | **ARTIC** | **GPAS** |  |
| 1 | Illumina | 21JDelta | 21JDelta | AY.98 | AY.98.1 | Delta (B.1.617.2-like) | Delta (B.1.617.2-like) | 93 |
| 1 | ONT | 21JDelta | 21JDelta | AY.98 | AY.98 | Delta (B.1.617.2-like) | Delta (B.1.617.2-like) | 92 |
| 2 | Illumina | 21JDelta | 21JDelta | AY.5 | AY.5 | Delta (B.1.617.2-like) | Delta (B.1.617.2-like) | 99 |
| 2 | ONT | 21JDelta | 21JDelta | AY.5 | AY.5.4 | Delta (B.1.617.2-like) | Delta (B.1.617.2-like) | 95 |
| 3 | Illumina | 21JDelta | 21JDelta | AY.4 | AY.44 | Delta (AY.4-like) | Delta (B.1.617.2-like) | 77 |
| 3 | ONT | 21JDelta | 21JDelta | AY.4 | AY.4 | Delta (AY.4-like) | Delta (AY.4-like) | 91 |
| 4 | Illumina | 21JDelta | 21JDelta | AY.4 | AY.4 | Delta (AY.4-like) | Delta (AY.4-like) | 99 |
| 4 | ONT | 21JDelta | 21JDelta | B.1.617.2 | AY.39 | Delta (B.1.617.2-like) | Delta (B.1.617.2-like) | 74 |
| 5 | Illumina | 21JDelta | 21JDelta | B.1.617.2 | None | Delta (B.1.617.2-like) | None | 55 |
| 5 | ONT | 21JDelta | 21JDelta | AY.4.2 | AY.4.2 | Delta (AY.4.2-like) | Delta (AY.4.2-like) | 94 |
| 6 | Illumina | 21JDelta | 21JDelta | AY.4 | None | Delta (AY.4-like) | None | 63 |
| 6 | ONT | 21JDelta | 21JDelta | AY.4 | AY.4 | Delta (AY.4-like) | Delta (AY.4-like) | 90 |
| 7 | Illumina | 21JDelta | 21JDelta | AY.4 | None | Delta (AY.4-like) | None | 59 |
| 7 | ONT | 21JDelta | 21JDelta | B.1.617.2 | AY.122 | Delta (B.1.617.2-like) | Delta (B.1.617.2-like) | 68 |
| 8 | Illumina | 21JDelta | 21JDelta | AY.4 | None | Delta (AY.4-like) | None | 60 |
| 8 | ONT | 21JDelta | 21JDelta | AY.4 | AY.4 | Delta (AY.4-like) | Delta (AY.4-like) | 60 |

* Out of a total of 99 amplicons

#### 2.2 TAP validation

TAP was upgraded from GPAS in 2024, with the following differences:

1. Human read removal: subtractive removal of human sequences in TAP (Hostile), rather than exclusive retention of SARS-CoV-2 sequences in GPAS (ReaditandKeep [15]);
2. Genome assembly: Viridian version 1.3.1 in TAP, and Viridian version 0.1.0 in GPAS;
3. Lineage assignment: Pangolin version 4.3.1 in TAP, and Pangolin version 4.1.1 in GPAS;
4. Relatedness: FindNeighbour5 in TAP, and FindNeighbour4 in GPAS [11].

Below, we describe a series of steps to assess concordance of TAP with GPAS.

**2.2.1	Lineage call concordance with a truth set**

Concordance of lineage calls made by TAP and GPAS was compared for the 12 cultured ‘truth’ samples described in section 2.1.2 (sequenced three-ways, excluding the SISPA runs). This produced 100% concordance of lineage calls (using the same Pango lineage definitions at the time of the creation of the ‘truth’ samples).

**2.2.2	Nucleotide call concordance with GPAS**

Concordance of nucleotide calls made by TAP and GPAS was compared using the 5,432 sequences from the current study. Comparing 3,862 assemblies that were assembled in both GPAS and TAP, there were 64 discordant sites (excluding non-ACGT sites), which corresponds to a discordance rate of 0.59/1,000,000 nucleotides. When restricting the comparison to 1,448 assemblies with no amplicon dropouts in both pipelines, this rate was even lower, with 12 discordant sites corresponding to a discordance rate of 0.28/1,000,000 nucleotides.

|  | **Total nucleotide sites** | **Discordant sites** | **Discordant rate per 1,000,000 sites** |
| --- | --- | --- | --- |
| No amplicon dropouts (n=1,448) | 42,875,563 | 12 | 0.28 |
| With amplicon dropouts (n=2,414) | 65,459,218 | 52 | 0.79 |
| **Total (n=3,862)** | **108,334,781** | **64** | **0.59** |

**2.2.3	Lineage call concordance with GPAS**

Finally, concordance of lineage calls made by TAP and GPAS was compared using the 5,432 sequences from the current study. Initial comparison identified 24% (1,323/5,432) discrepancies, of which the majority (84%, 1,108/1,323) resolved on rolling back Pangolin versions to be like-for-like. A further 14% (180/1,323) were resolved on rolling back Viridian versions to be like-for-like; direct comparison of genome assemblies showed that this is likely due to the newer Viridian version having greater mean coverage and differences in non-ACGTN ambiguity calls; all differences in lineage calls were in any case closely related, with a mean SNP difference of 1.8 (SD: 2.4, range 0-100). The remaining 2.6% (35/1,323) originated from one batch from one submitting site, which were unassembled in GPAS but assembled and assigned lineages in TAP. This discrepancy was determined to be caused by a batch processing issue in the legacy GPAS platform at the time of sample upload. Overall, lineage concordance between GPAS and TAP, accounting for component version upgrades, was 99.4% (5,397/5,432).

### 3 Supplementary Appendix C: Tables and Figures

#### 3.1 Supplementary Table 1

Summary.csv file with the output from TAP.

#### 3.2 Supplementary Table 2

The table below shows all unique isolate pairs at each SNV distance (0, 1, 2, 3, or >3), for the three major lineages (BA.2, BA.4, BA.5). Concordant sites indicate that both isolates were from the same site, and discordant sites indicate different sites. The proportion of discordance with 95% confidence intervals is shown.

| **SNV distance** | **Unique isolate pairs (n)** | | |  | **Proportion of discordance (95% CI)** |
| --- | --- | --- | --- | --- | --- |
|  | **Total** | **Concordant sites** | **Discordant sites** |  |  |
| **BA2** |  |  |  |  |  |
| **0** | 216 | 170 | 46 |  | 0.213 (0.158 – 0.268) |
| **1** | 630 | 354 | 276 |  | 0.438 (0.399 – 0.477) |
| **2** | 1921 | 1055 | 866 |  | 0.451 (0.429 – 0.473) |
| **3** | 3954 | 2211 | 1743 |  | 0.441 (0.425 – 0.456) |
| **>3** | 693927 | 125132 | 568795 |  | 0.820 (0.819 – 0.821) |
| **BA4** |  |  |  |  |  |
| **0** | 3494 | 3340 | 154 |  | 0.044 (0.037 – 0.051) |
| **1** | 9003 | 8463 | 540 |  | 0.060 (0.055 – 0.065) |
| **2** | 15494 | 13989 | 1505 |  | 0.097 (0.092 – 0.102) |
| **3** | 22340 | 19060 | 3280 |  | 0.147 (0.142 – 0.151) |
| **>3** | 1942202 | 329942 | 1612260 |  | 0.830 (0.830 – 0.831) |
| **BA5** |  |  |  |  |  |
| **0** | 2517 | 2082 | 435 |  | 0.173 (0.158 – 0.188) |
| **1** | 6017 | 2853 | 3164 |  | 0.526 (0.513 – 0.538) |
| **2** | 15343 | 6038 | 9305 |  | 0.606 (0.599 – 0.614) |
| **3** | 30268 | 10867 | 19401 |  | 0.641 (0.636 – 0.646) |
| **>3** | 2377475 | 728597 | 1648878 |  | 0.694 (0.693 – 0.694) |

#### 3.3 Supplementary Figure 1

Distribution of processing time (in minutes) for each sample in the GPAS pipeline according to submitting site for all samples.

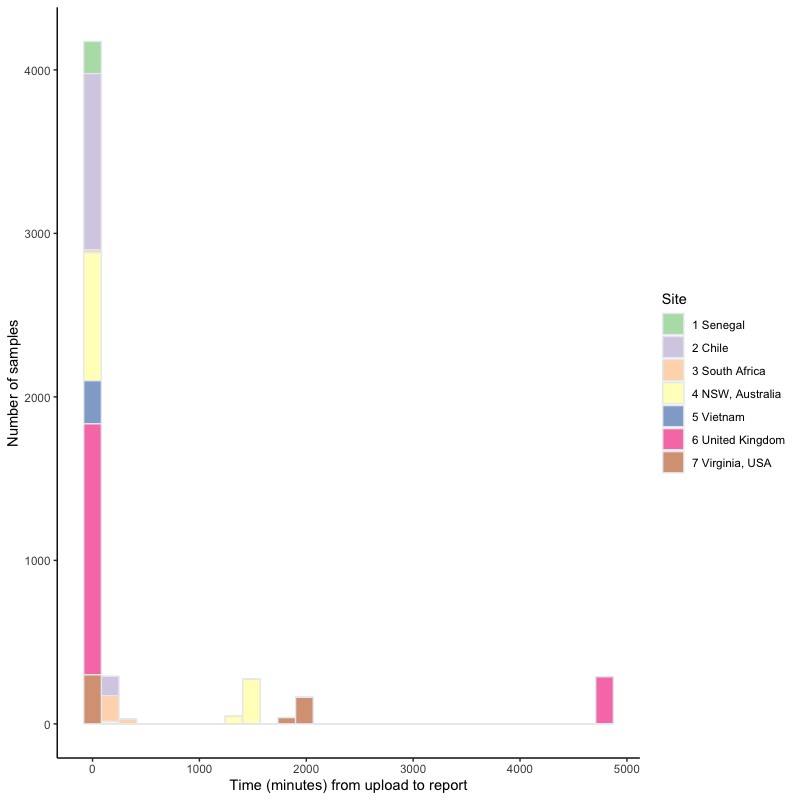

#### 3.4 Supplementary Figure 2

Processing time in TAP from upload to result for representative batches of 10, 50, 100 and 500 Illumina or ONT samples. Times are shown for both the total run time per batch, and median time per sample.

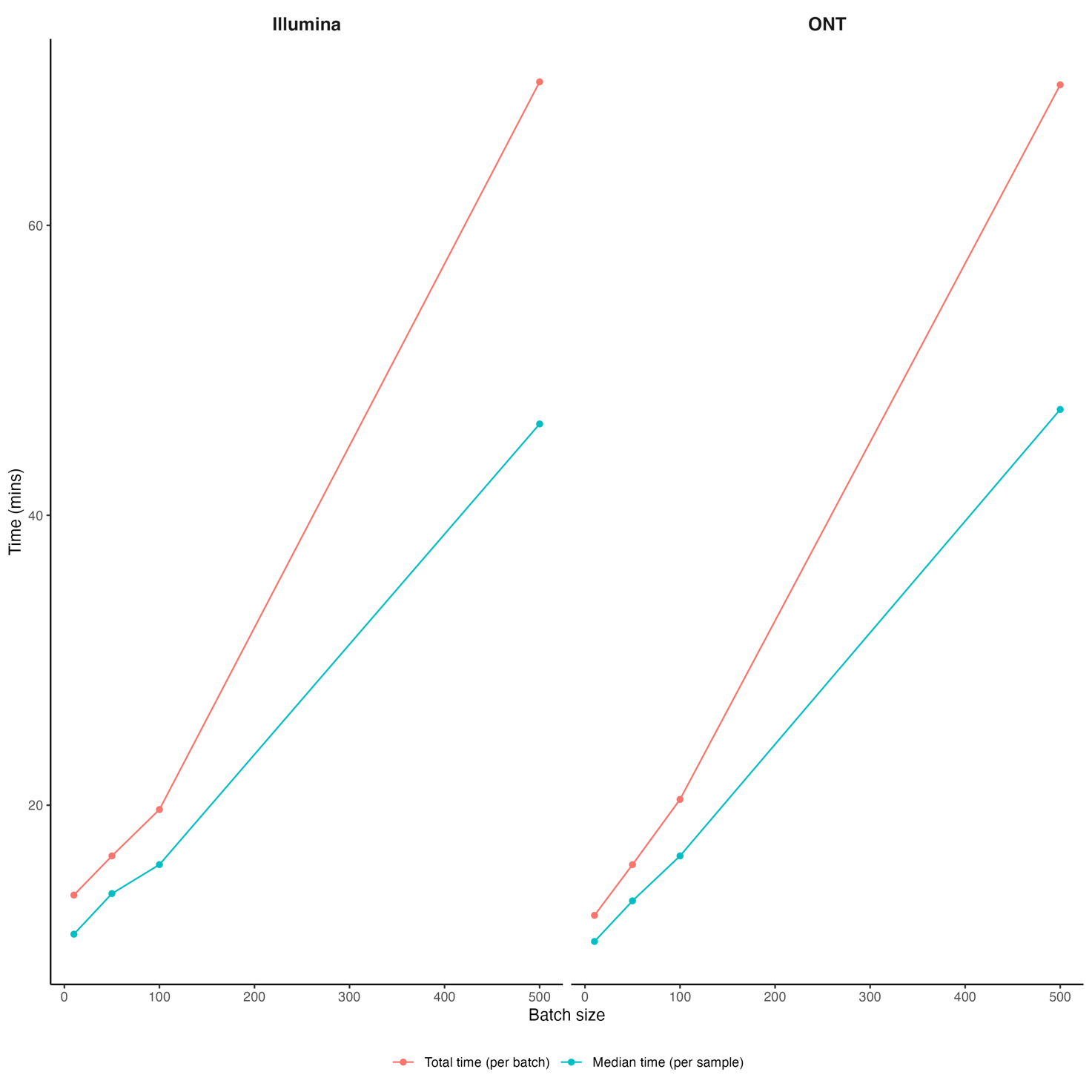

**References**

1.	Westhead J, Baker CS, Brouard M, Colpus M, Constantinides B, Hall A, et al. Enhancement and validation of the antibiotic resistance prediction performance of a cloud-based genetics processing platform for Mycobacteria. bioRxiv. 2024:2024.11.08.622466.

2.	Constantinides B, Hunt M, Crook DW. Hostile: accurate decontamination of microbial host sequences. Bioinformatics. 2023;39(12).

3.	Li H. Minimap2: pairwise alignment for nucleotide sequences. Bioinformatics. 2018;34(18):3094-100.

4.	Langmead B, Salzberg SL. Fast gapped-read alignment with Bowtie 2. Nat Methods. 2012;9(4):357-9.

5.	Danecek P, Bonfield JK, Liddle J, Marshall J, Ohan V, Pollard MO, et al. Twelve years of SAMtools and BCFtools. Gigascience. 2021;10(2).

6.	Wood DE, Lu J, Langmead B. Improved metagenomic analysis with Kraken 2. Genome Biol. 2019;20(1):257.

7.	Hunt M, Hinrichs AS, Anderson D, Karim L, Dearlove BL, Knaggs J, et al. Addressing pandemic-wide systematic errors in the SARS-CoV-2 phylogeny. bioRxiv. 2024:2024.04.29.591666.

8.	O'Toole A, Scher E, Underwood A, Jackson B, Hill V, McCrone JT, et al. Assignment of epidemiological lineages in an emerging pandemic using the pangolin tool. Virus Evol. 2021;7(2):veab064.

9.	McCrone JT, Hill V, Bajaj S, Pena RE, Lambert BC, Inward R, et al. Context-specific emergence and growth of the SARS-CoV-2 Delta variant. Nature. 2022;610(7930):154-60.

10.	FindNeighbour5 [Available from: <https://github.com/oxfordmmm/FN5>.

11.	FindNeighbour4 [Available from: <https://github.com/davidhwyllie/findNeighbour4/blob/master/reference/covid_exclusion.vcf>.

12.	ncov2019-artic-nf [Available from: <https://github.com/connor-lab/ncov2019-artic-nf>

13.	Thielen PM, Wohl S, Mehoke T, Ramakrishnan S, Kirsche M, Falade-Nwulia O, et al. Genomic diversity of SARS-CoV-2 during early introduction into the Baltimore-Washington metropolitan area. JCI Insight. 2021;6(6).

14.	Constantinides B, Webster H, Rodger G, Hunt M, Supasa P, Dejnirattisai W, et al. A diverse reference set of cultured SARS-CoV-2 genomes sequenced using various amplification methods and instrument platforms 2024 [Available from: <https://www.ebi.ac.uk/biostudies/studies/S-BSST1334>.

15.	Hunt M, Swann J, Constantinides B, Fowler PW, Iqbal Z. ReadItAndKeep: rapid decontamination of SARS-CoV-2 sequencing reads. Bioinformatics. 2022;38(12):3291-3.
